# United States’ qualifying conditions compared to evidence of the 2017 National Academy of Sciences Report

**DOI:** 10.1101/2023.05.01.23289286

**Authors:** Elena L. Stains, Amy L. Kennalley, Maria Tian, Kevin F. Boehnke, Chadd K. Kraus, Brian J. Piper

## Abstract

**Objective:** To compare the 2017 National Academies of Sciences, Engineering, and Medicine (NAS) report to state medical cannabis (MC) laws defining approved qualifying conditions (QC) from 2017 to 2024 and to determine if there exist gaps in evidence-based decision making.

**Methods:** The 2017 NAS report assessed therapeutic evidence for over twenty medical conditions treated with MC. We identified the QCs of 38 states (including Washington, D.C.) where MC was legal in 2024. We also identified the QCs that these states used in 2017. QCs were then categorized based on NAS-established level of evidence: substantial/conclusive evidence of effectiveness, moderate evidence of effectiveness, limited evidence of effectiveness, limited evidence of ineffectiveness, and no/insufficient evidence to support or refute effectiveness. This study was completed between January 31, 2023 through May 20, 2024.

**Results:** Most states listed at least one QC with substantial evidence—80.0% of states in 2017 and 97.0% in 2024. However, in 2024 only 8.3% of the QCs on states’ QC lists met the standard of substantial evidence. Of the 20 most popular QCs in the country in 2017 and 2024, one only (chronic pain) was categorized by the NAS as having substantial evidence for effectiveness. However, seven (ALS, Alzheimer’s disease, epilepsy, glaucoma, Huntington’s disease, Parkinson’s disease, spastic spinal cord damage) were rated as either ineffective or insufficient evidence.

**Conclusion:** Most QCs lack evidence for use based on the 2017 NAS report. Many states recommend QCs with little evidence, such as amyotrophic lateral sclerosis (ALS), or even those for which MC is ineffective, like depression. There have been insufficient updates to QCs since the NAS report. These findings highlight a disparity between state-level MC recommendations and the evidence to support them.

## Introduction

As of 2024, 37 states and the District of Columbia, representing 73% of the United States (US) population, have legislation for medical cannabis (MC) (1). Each state determines the qualifying conditions (QCs) that allow patients to receive recommendation for MC by providers. QCs range widely between states, and commonly include conditions such as chronic pain, anxiety, and post-traumatic stress disorder (PTSD) (2). In 2019-2020, 2.5% of Americans reported using cannabis for medical needs, compared to 1.2% in 2013-2014, representing a 12.9% annual increase (1).

Like any pharmaceutical agent, there are potential clinical benefits and harms associated with MC and research into therapeutic uses of MC continues to evolve (3,4). This presents a potential public health challenge, as the societal and political acceptance of the drug might be moving faster than scientific understanding. One consideration for patients is cost—MC is not covered by Medicare, Medicaid, or private insurance and therefore is an out-of-pocket expense for most (although Pennsylvania recently implemented a Medical Marijuana Assistance Program to help pay the cost of MC and card fees for eligible patients) (5). One study conducted among medical cannabis dispensary patients in New England reported that many were spending over two-thousand dollars per year (3). There is also concern about drug interactions, particularly involving CYP3A4, with the prescription formulations of tetrahydrocannabinol (THC) and cannabidiol (6). While lethal overdose from cannabis is extremely rare, there are rare but serious case-reports of fatalities involving cardiac events, and considerable societal and human costs associated with cannabis hyperemesis syndrome (7–9).

The National Academies of Sciences, Engineering, and Medicine (NAS) published a report in 2017 on the evidence, or lack thereof, for the therapeutic effects of MC for over twenty conditions (10). This landmark report involved reviewing more than 10,700 abstracts and has the potential to be a guide for states looking for a scientific guidance for approving QCs (10). However, it has not always been used for this process, with states reportedly making the decision to include QCs by also incorporating expert opinion, evidence, and public wishes (11,12). The state of Delaware allows citizens to petition to add QCs and approves such conditions based on two main factors: “(1) the medical condition or treatment is debilitating and (2) marijuana is more likely than not to have the potential to be beneficial to treat or alleviate the debilitation associated with the medical condition or treatment (13).”

The process that states have applied thus far has resulted in a gap between the QCs recommended by states and the evidence to support MC use for those conditions. For example, of Pennsylvania’s 24 QCs in 2024, the NAS indicates that only two (8.3%), “severe chronic or intractable pain” and “neuropathies,” had conclusive or substantial evidence that cannabinoids were effective while 25%, including Amyotrophic Lateral Sclerosis, glaucoma, epilepsy, Huntington’s disease, Parkinson’s disease, and opioid use disorder, were categorized as conditions for which cannabinoids were not shown to be effective (14). Other studies have incorporated the 2017 NAS report to comment on public policy (14,15), but none have analyzed how QCs of every state compare to the evidence in the NAS report.

In this report, we detail the past and present QCs of all states in which MC was legal from 2017 to 2024. We map the evidence of each QC and exhibit that QCs with substantial evidence on average make up less than a tenth of each states’ list. Our analysis shows that not only do states’ lists of QCs exhibit a discordance with the available evidence, but states have not progressed to high-evidence QC lists over time. This revelation exposes a deficit in evidence-based recommendations for MC use in public policy. Through the findings in this research, we can guide safer, more informed public health legislation for patients.

## Methods

### Procedures

QCs were collected for each of the 37 states and the District of Columbia where MC was legal in the United States as of April 2023 when analyses began. Data for those states that had MC policy in April 2023 was updated for 2024, but states that created MC policy since April 2023 were not added to our analysis. Two states, Kentucky and Georgia, had legislation to allow a low-THC form of MC for certain conditions, but were excluded from this analysis. For ease of reporting, we classified the District of the Columbia as a state, making 38 “states” with MC in 2024. This information was verified by comparison to each state’s medical cannabis program (see: Supplemental Appendix for references). We used an internet archive tool to collect the QCs for each state in 2017 (16). Of the 38 states with legalized MC in 2024, 31 had legalized MC in 2017. The NAS report was published in 2017, making that the first year of our data collection (10). Our objective was to compare states’ QCs at the time of publication of the NAS report to subsequent years and to identify whether states updated their QC lists to align with the most up-to-date evidence.

The NAS report was used as our “gold standard” of evidence because it is a comprehensive review of available evidence for twenty of the most popular QCs in the country. The NAS committee that created the report considered over 10,000 systematic reviews and primary studies. The final report contains over 400 pages of summary of evidence for both the benefits and harms of MC (10).

Ten states (CA, DC, LA, ME, MD, MA, MO, NY, OK, VA) in 2024 included “blanket statements” that permitted provider discretion to recommend MC for any condition that they deemed necessary. These statements were not counted as QCs and were not included in a state’s total number of QCs. Five states (DC, ME, NY, OK, VA) had no QCs whatsoever, allowing full discretion to certifying providers.

Each condition was divided into the categories established by the NAS report: conclusive/substantial evidence of effectiveness (e.g. chronic pain), moderate evidence of effectiveness (e.g. improved sleep outcomes for certain conditions), limited evidence of effectiveness (e.g. PTSD), limited evidence of ineffectiveness (e.g. glaucoma), and no/insufficient evidence to support or refute effectiveness (e.g. epilepsy) (10) (Table 1A).

**Table 1.** Categories of evidence established by the 2017 National Academies of Sciences, Engineering, and Medicine (NAS) report. (a). Common state qualifying conditions for medical cannabis that were deemed “partial” due to broad wording, compared with evidence found by NAS (b) (10).

| **National Academies of Sciences categories of evidence** | **Conditions/symptoms** |
| --- | --- |
| Conclusive or Substantial evidence of effectiveness | For the treatment of chronic pain in adults<br>As antiemetics in the treatment of chemotherapy-induced nausea and vomiting<br>For improving patient-reported multiple sclerosis spasticity symptoms |
| Moderate evidence of effectiveness | Improving short-term sleep outcomes in individuals with sleep disturbance associated with obstructive sleep apnea syndrome, fibromyalgia, chronic pain, and multiple sclerosis |
| Limited evidence of effectiveness | Increasing appetite and decreasing weight loss associated with HIV/AIDS<br>Improving clinician-measured multiple sclerosis spasticity symptoms<br>Improving symptoms of Tourette syndrome<br>Improving anxiety symptoms, as assessed by a public speaking test, in individuals with social anxiety disorders<br>Improving symptoms of posttraumatic stress disorder |
| Limited evidence of a statistical association | Better outcomes (i.e., mortality, disability) after a traumatic brain injury or intracranial hemorrhage |
| Limited evidence of ineffectiveness | Improving symptoms associated with dementia<br>Improving intraocular pressure associated with glaucoma<br>Reducing depressive symptoms in individuals with chronic pain or multiple sclerosis |
| No/insufficient evidence to support or refute effectiveness | Cancers, including glioma<br>Cancer-associated anorexia cachexia syndrome and anorexia nervosa<br>Symptoms of irritable bowel syndrome |
|  | Epilepsy |
|  | Spasticity in patients with paralysis due to spinal cord injury |
|  | Symptoms associated with amyotrophic lateral sclerosis |
|  | Chorea and certain neuropsychiatric symptoms associated with Huntington's disease |
|  | Motor system symptoms associated with Parkinson's disease or the levodopa-induced dyskinesia |
|  | Dystonia |
|  | Achieving abstinence in the use of addictive substances |
|  | Mental health outcomes in individuals with schizophrenia or schizophreniform psychosis |

b.
| Qualifying Condition | NAS evidence related to this condition |
| --- | --- |
| Anxiety | Limited evidence for: anxiety symptoms in those with social anxiety |
| Cachexia | Limited evidence for: increased appetite and decreased weight loss in HIV/AIDS. Insufficient Evidence for: anorexia/cachexia syndrome in cancer |
| Cancer | Conclusive/substantial evidence of effectiveness for: chronic pain in cancer, chemotherapy-induced nausea and vomiting. Insufficient evidence for: cancer regression, cancer-associated anorexia/cachexia syndrome. |
| HIV or AIDS | Limited evidence of effectiveness for: increased appetite and decreased weight loss in HIV/AIDS |
| Multiple Sclerosis | Substantial evidence for: patient-reported MS spasticity, chronic pain due to MS.<br>Moderate evidence for: improved short term sleep outcomes for MS.<br>Limited evidence for: clinician-measured MS spasticity.<br>Limited evidence of ineffectiveness for: depressive symptoms due to MS. |
| Nausea | Conclusive evidence for: chemotherapy-induced nausea and vomiting. |
| Seizure | Insufficient evidence for: Epilepsy |
| Severe and persistent muscle spasms | Substantial evidence for: patient-reported MS spasticity.<br>Limited evidence for: clinician-measured MS spasticity.<br>Insufficient evidence for: spasticity due to paralysis due to spinal cord injury, dystonia, Parkinson's disease motor system symptoms or levodopa-induced dyskinesia. |

QCs that only partially fit into the NAS-established categories, when taken exactly as written, were categorized as “partial” (Table 1B). For example, 93.4% of states list “cancer” as a QC and although there are several applications of MC for cancer listed in the NAS report (e.g. antiemetic for chemo induced nausea/vomiting, cancer-associated anorexia cachexia syndrome, chronic pain associated with cancer) each had a different level of evidence. Because it was broad, cancer and other broad QCs were categorized as partial. As any other QC, each “partial” QC counted as one in the QC count.

Many states listed QCs that were not studied in the NAS report (e.g. Crohn’s disease) and these were categorized as “N/A.”

### Data analysis

Chi-square was completed with GraphPad Prism to analyze differences (p < 0.05) in QCs of each category across years 2017 to 2024 (17) (Supplemental Table 1). Figures were prepared with Prism (v10.0) and heatmaps were constructed using HeatMapper (17).

## Results

### Summary of QCs, 2017 and 2024

Thirty-one states (including the District of Columbia) had legalized MC in 2017. The number of QCs varied five-fold between states (Min = 8 in Alaska, Colorado, Maryland, and Massachusetts, Max = 40 in Illinois) with a mean of 14.7 (Fig 1).

**Figure 1.**
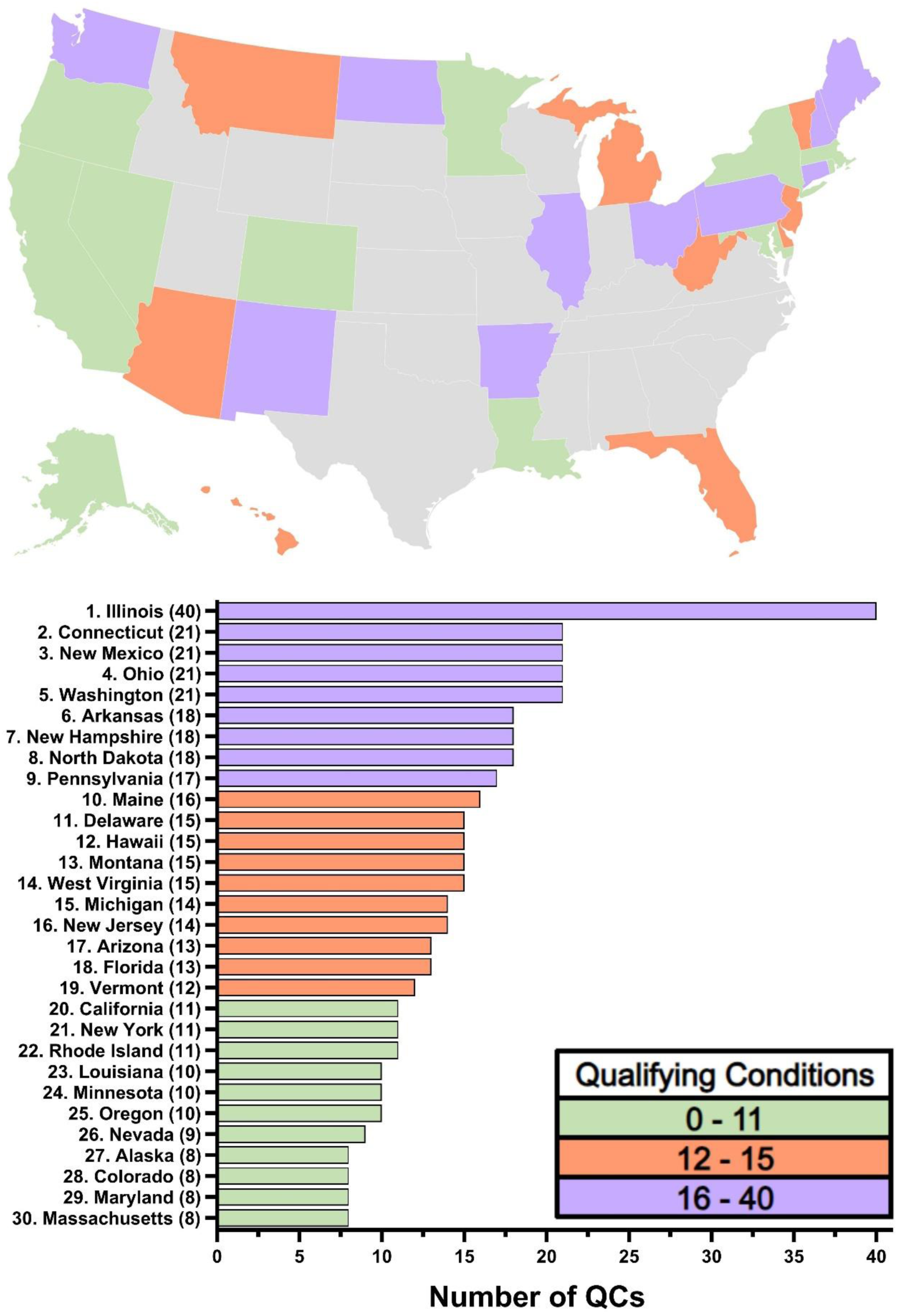
Number of approved Qualifying Conditions (QC) for medical cannabis (MC) per state in 2017. The District of Columbia was not displayed due to lack of QC list. States that did not have medical cannabis in 2017 are shown in grey.

In 2024, 38 states had legalized MC. The average number of QCs in a state was 18.7. Overall, states in the western US had fewer QCs than those in the Midwest. There was a ten-fold difference in QCs between the state with the fewest (South Dakota, 5) and the most (Illinois, 52) (Fig 2). Ten states included with their list of QCs the ability of providers to recommend MC at their discretion–while five (District of Columbia, Maine, New York, Oklahoma, Virginia) left the decision to the discretion of the provider, requiring no QCs whatsoever.

**Figure 2.**
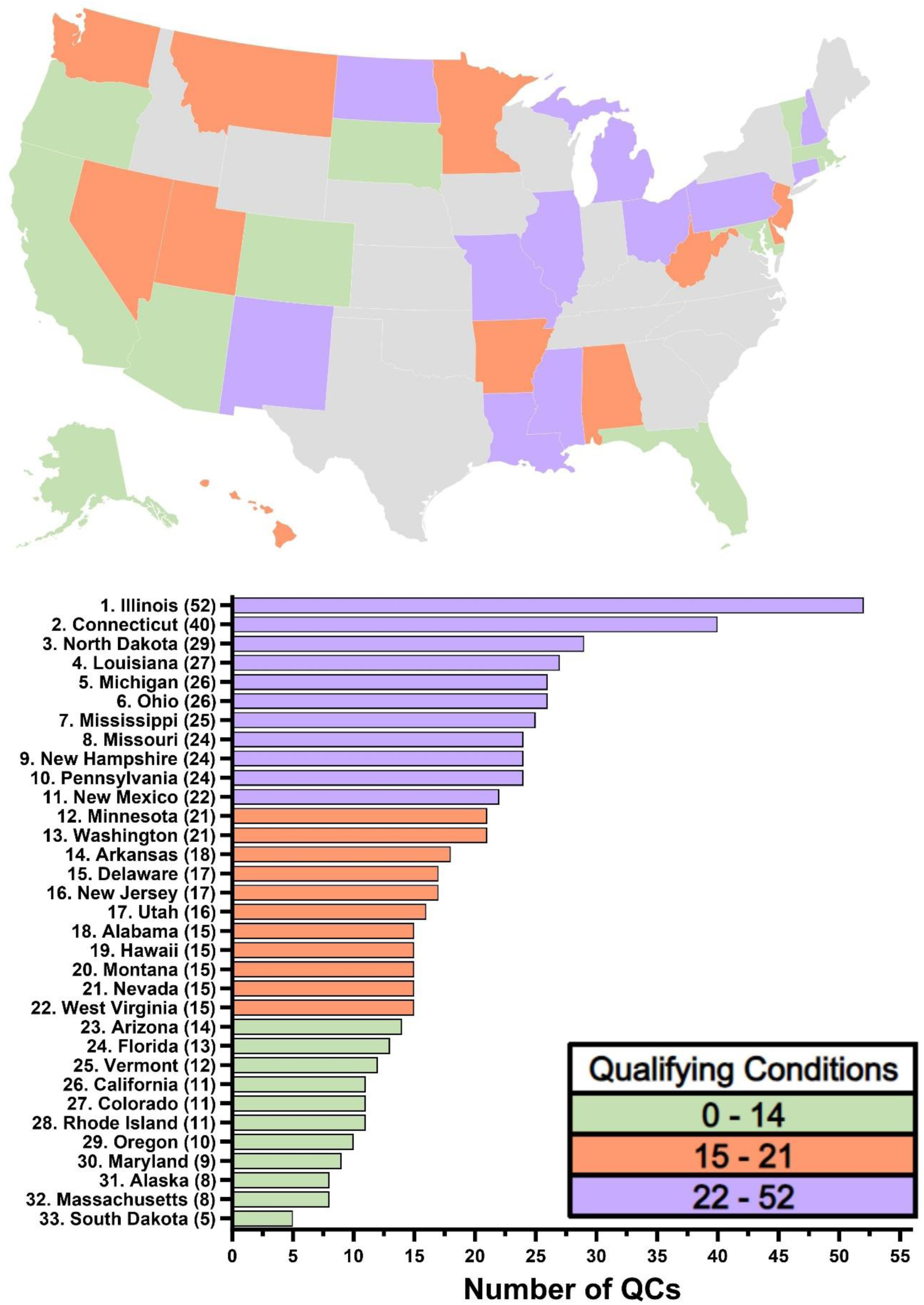
Number of approved Qualifying Conditions (QC) for medical cannabis (MC) per state in 2024. The District of Columbia, Maine, New York, Oklahoma, and Virginia were not displayed due to lack of QC list. States that did not have medical cannabis in 2024 are shown in grey.

### Evidence of QCs, 2017 and 2024

In 2017, most (80.0%) states with a QC list had at least one QC with substantial evidence (Fig 3A). However, only 6.8% of the country’s QCs met the standard of substantial evidence. In contrast, 90.0% of states listed one or more QCs with limited evidence of ineffectiveness (Fig 3B).

**Figure 3.**
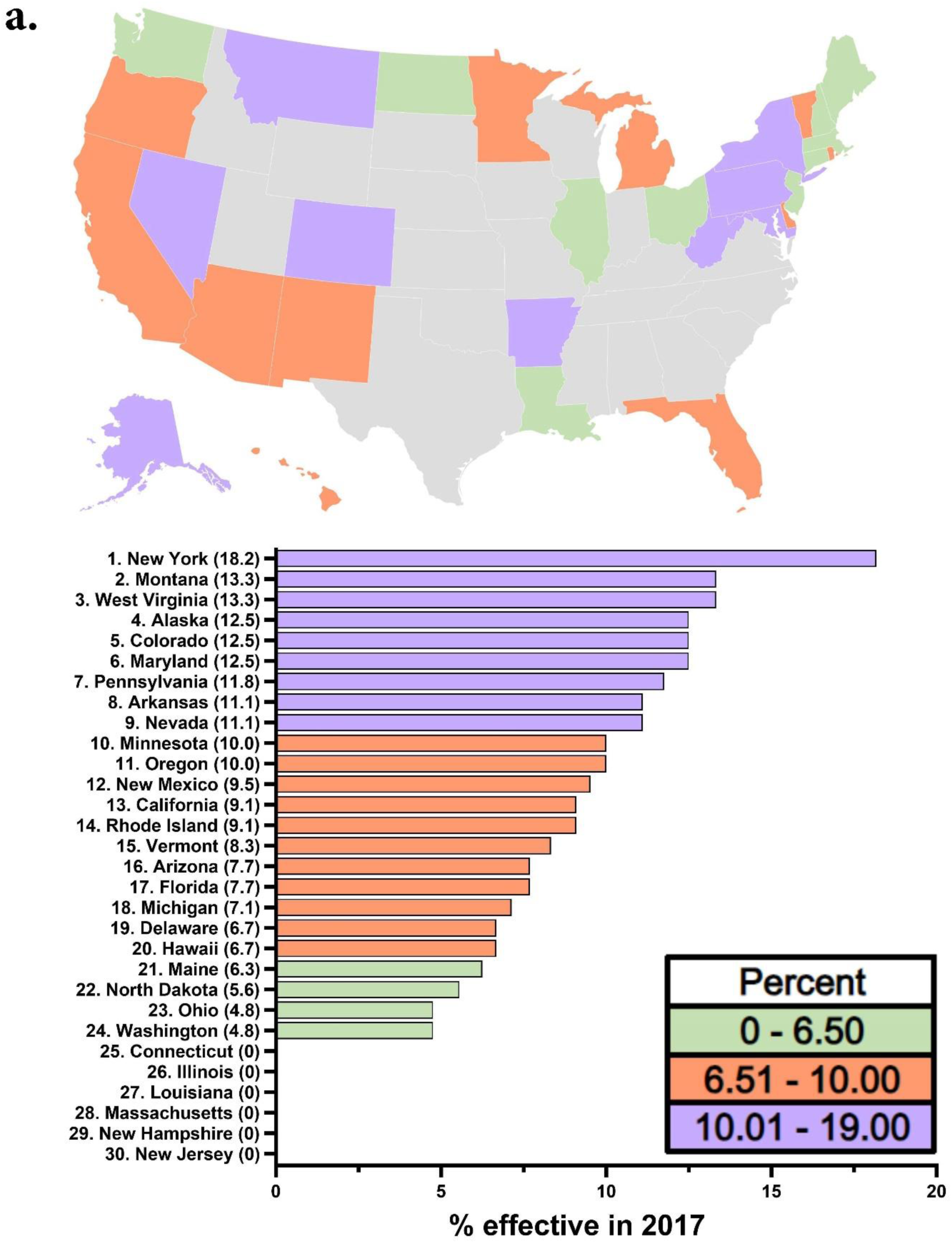
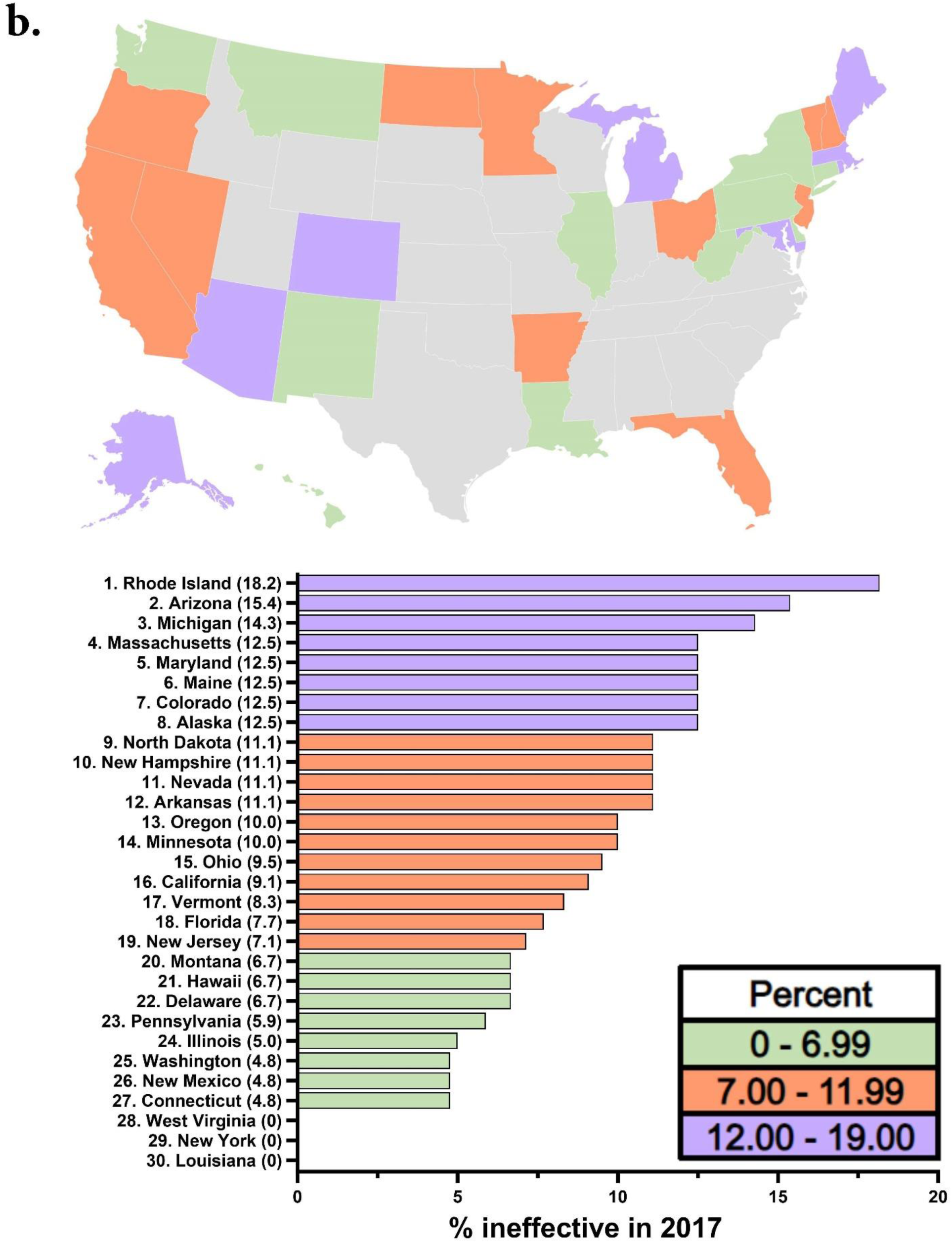
Percent of each state’s qualifying conditions (QC) that had substantial evidence of effectiveness (a) and limited evidence of ineffectiveness (b) in 2017 according to the National Academy of Sciences, Engineering, and Medicine (NAS). The District of Columbia was not displayed due to lack of QC list. States that did not have medical cannabis in 2017 are shown in grey.

In 2024, 97.0% states with a QC list had at least one QC with substantial evidence of effectiveness (Fig 4A), but only 8.3% of states’ QCs met the standard of substantial evidence. Ninety-one percent of states listed one or more QCs with limited evidence of ineffectiveness (Fig 4B), most commonly glaucoma.

**Figure 4.**
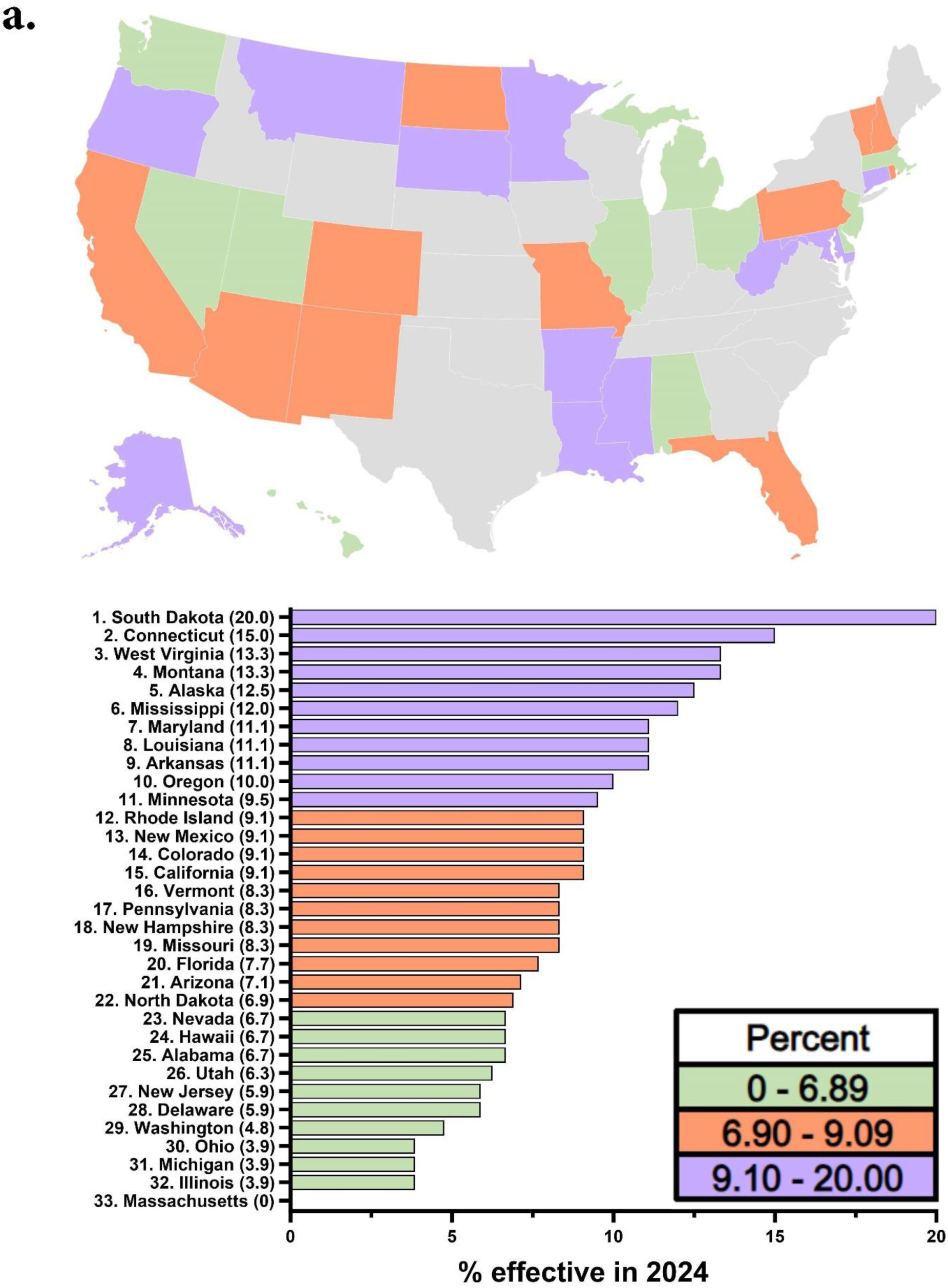
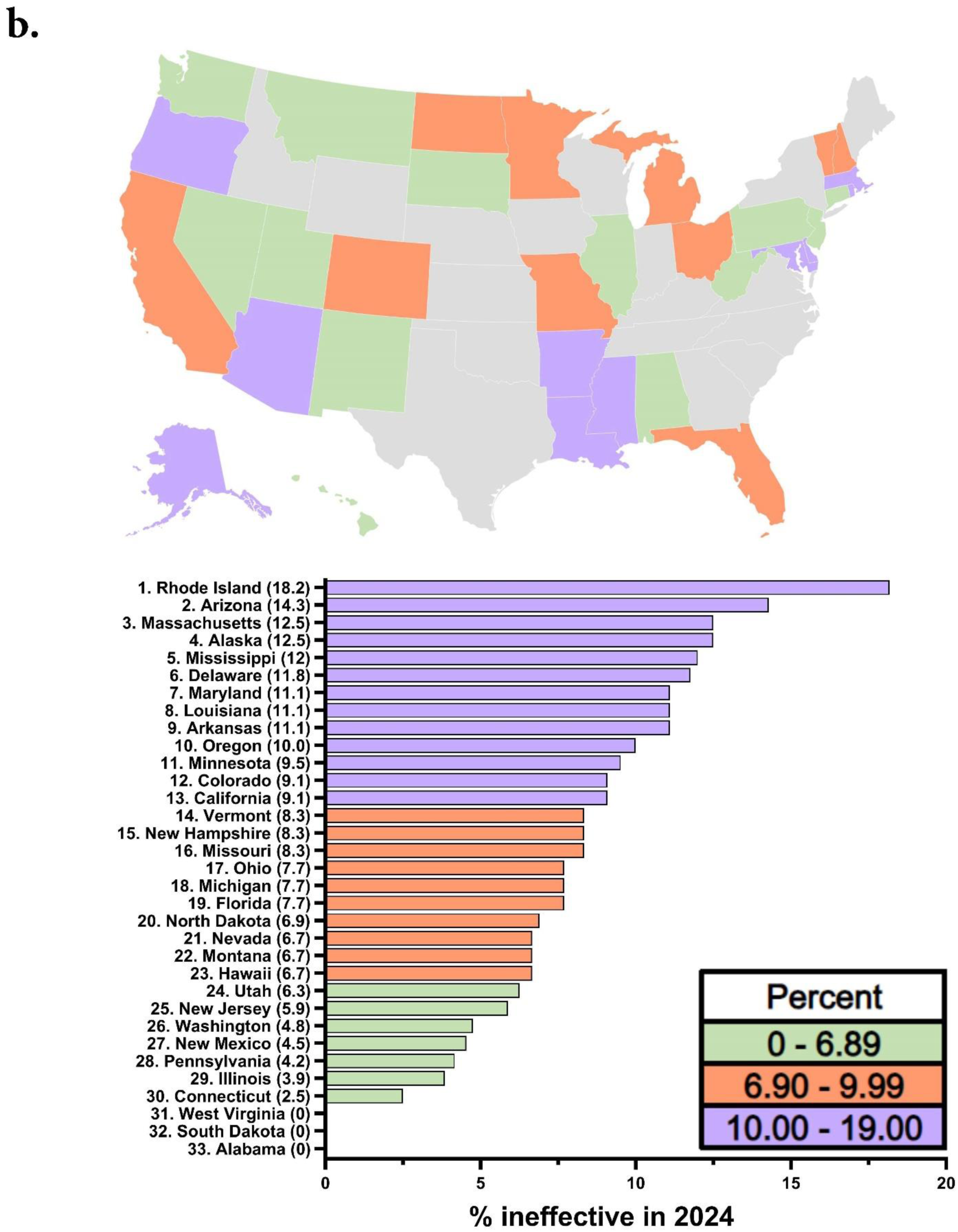
Percent of each state’s qualifying conditions for medical cannabis that had substantial evidence of effectiveness (a) and limited evidence of ineffectiveness (b) in 2024 according to the National Academy of Sciences, Engineering, and Medicine (NAS) (10). The District of Columbia, Maine, New York, Oklahoma, and Virginia were not displayed due to lack of QC list. States that did not have medical cannabis in 2023 are shown in grey.

In 2024, the majority (90.9%) of states had at least one QC that was not in the NAS report. Twenty-eight percent of the country’s QCs were not included in the NAS report, and 37.2% were a partial fit.

### Changes to QCs since 2017

Of the states with MC in 2017, 20.0% listed a higher percentage of QCs with substantial evidence in 2024, but another 33.3% listed a lower percentage. Half (46.7%) recommended a lower percentage of ineffective QCs by 2023, and only 6.7% recommended a higher percentage (Fig 5).

**Figure 5.**
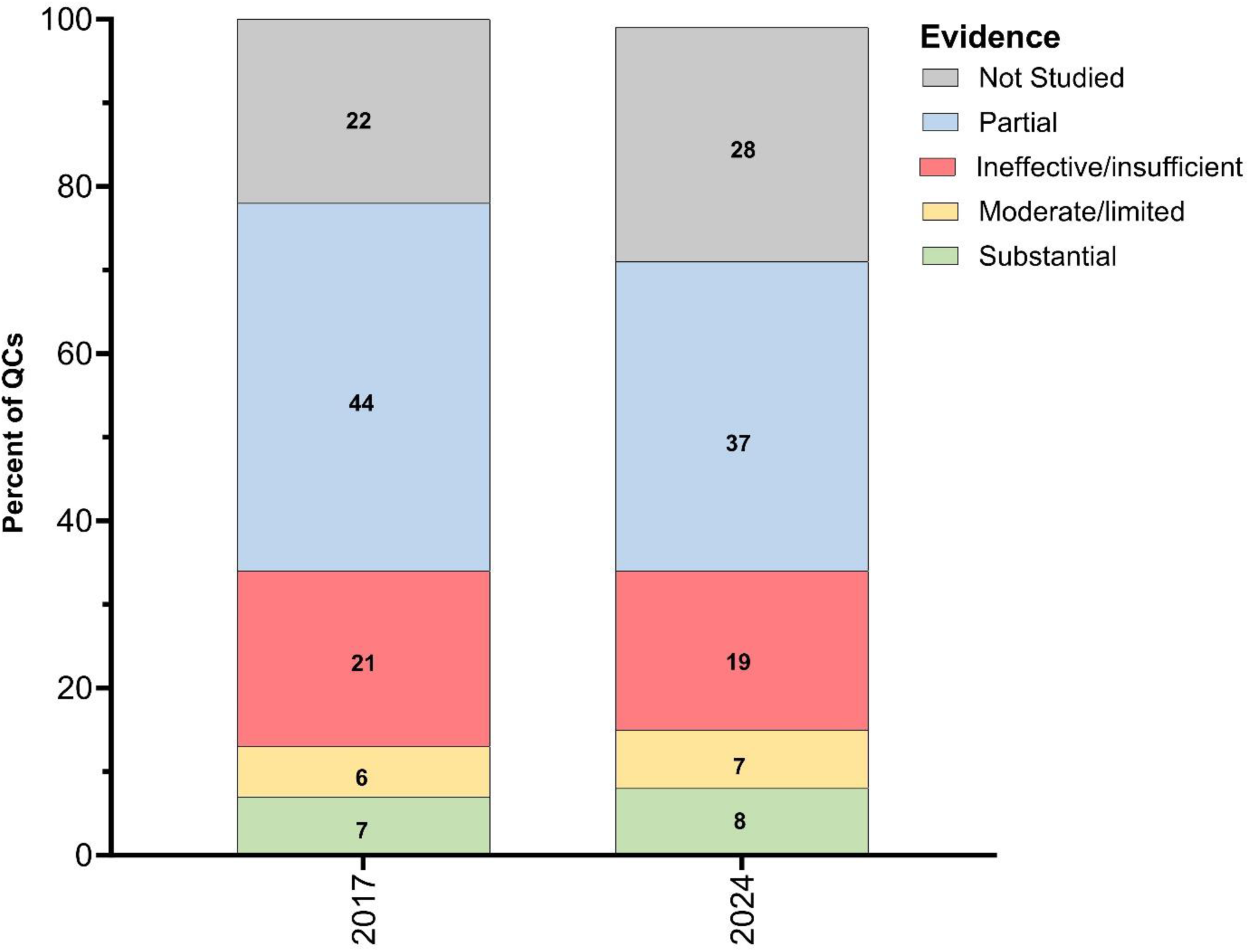
Number of total qualifying conditions (QC) in all states in 2017 and 2024. Each year is divided into and color-coded by percent of QCs in each of the National Academy of Sciences (NAS) established categories. States that had no QCs, only blanket statements (District of Columbia, Maine, New York, Oklahoma, and Virginia), were excluded.

In a chi-squared analysis, we found a significant increase of those QCs not studied by the NAS report. These QCs increased from 97 in 2017 to 171 in 2024. QCs labeled “partial” significantly increased from 195 to 229. There was no significant difference between the number of QCs with substantial evidence between 2017 and 2024 (Supplemental Table 1). This was also true of the other categories of evidence (moderate/limited evidence, evidence of ineffectiveness, and insufficient evidence). Of the 20 most popular QCs in the country in 2017 and 2024, one only (chronic pain) was categorized by the NAS as having substantial evidence for effectiveness (10). However, seven (ALS, Alzheimer’s disease, epilepsy, glaucoma, Huntington’s disease, Parkinson’s disease, spastic spinal cord damage) were rated as either ineffective or insufficient evidence (Fig 6).

**Figure 6.**
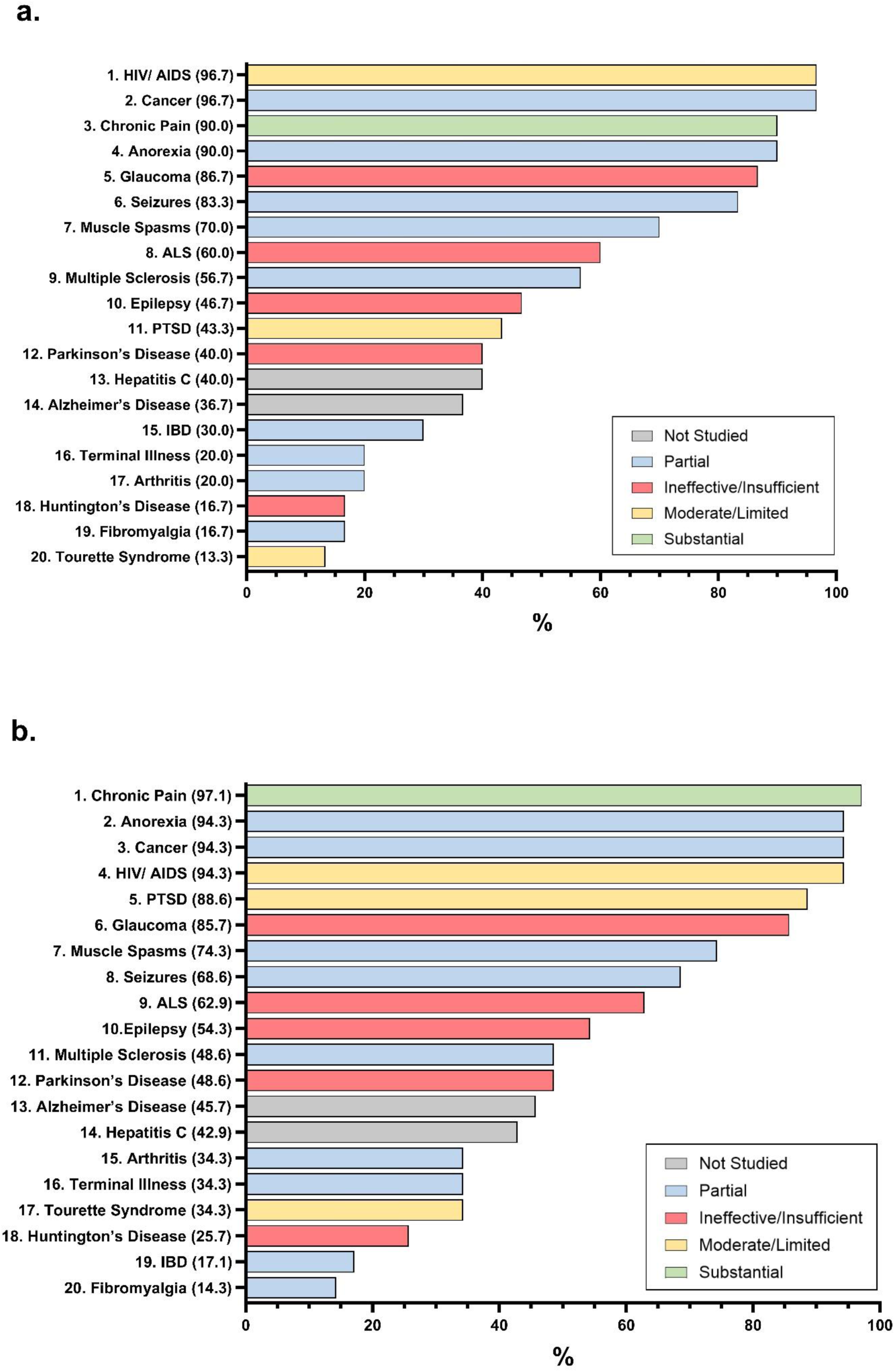
Percentage of states that include the mentioned QCs in their list in 2017 (a) and 2024 (b). Each QC is color-coded by its National Academy of Sciences (NAS) evidence rating.

## Discussion

In our analyses, we show that the majority of QCs (91.7%) that states use to qualify patients for MC recommendation do not align with evidence for benefit (10). Most states recommend QCs for which the effects of MC have not been well-studied–ALS, Parkinson’s, opioid dependence— or are known to some degree to be ineffective–Alzheimer’s, glaucoma, and Huntington’s (10). When comparing QCs from 2017 (the time of publication of the NAS report) to 2024, there are no data to suggest that states have updated their recommendations based on the evidence in the NAS report. In fact, based on our chi-squared analysis, the only types of QCs that have significantly increased are those not reported on by the NAS report, and those more vaguely titled QCs that we labeled partial.

Other sources of evidence-based medicine have also been ignored by states. For example, since 2009 the American Glaucoma Society has stated that MC is *not* recommended for glaucoma due to “its side effects and short duration of action, coupled with a lack of evidence that it use alters the course of glaucoma (18).” The Parkinson’s Foundation released the following consensus statement in 2020 on MC for Parkinson’s disease (PD): “Given the lack of any clear data supporting the use of cannabis in PD, the Foundation does not endorse their use for PD symptoms or to modify disease progression (19).” Despite these clear statements against MC from leading professional and advocacy organizations, many states continue to recommend MC for those diseases.

According to our data, states added QCs to their lists over time from all categories. However, there was no evidence that states revised and updated QC lists according to levels of available evidence. The non-significant results of the time analysis suggest that changes in the number of QCs with substantial evidence over time is more likely due to other, to be determined, factors besides an evidence-based alignment. The changes in QC categories over time (Fig 6) may suggest that states had decreased the percentage of ineffective QCs between 2017 and 2024. However, not one state removed QCs from their list during that period. In this way, states only lowered their percentage of QCs labeled ineffective by dilution.

Although most listed QCs do not have substantial evidence of effectiveness, according to a study of 19 states in 2022, the most commonly reported use for MC was chronic pain (20). Used by 48.4% of patients, chronic pain does fall under the category of substantial evidence (20). The second most common (14.2%), anxiety, was marked as partial in our analysis, with the NAS report citing “limited evidence of effectiveness for anxiety symptoms in individuals with social anxiety (10,20).” The third most common (13.0%), PTSD, had limited evidence of effectiveness (10,20).

States recommending QCs that are ineffective is a potential public health concern. First, the money patients spend on MC, which, according to one study, averaged 3,064 $US/year in 2018, may be better utilized on more evidence-based interventions (3). Second, the prescription formulations of THC and CBD, the most abundant components of the *Cannabis sativa* plant, have known drug interactions (6). Third, cannabis potentially possess adverse effects and safety concerns. States have been challenged to incorporate traffic safety laws into the growing landscape of legalized cannabis. Colorado law attempts to control driving under the influence of cannabis by creating a blood THC level (five nanograms of THC per milliliter of blood) over which drivers can be prosecuted for impaired driving, even though the National Institute of Justice found that body fluid levels of THC cannot accurately or reliably predict level of impairment (21,22). The NAS report includes evidence for harms of MC which found that smoking cannabis increases risk for respiratory symptoms and chronic bronchitis episodes (10). They also reported on substantial evidence of an association between cannabis use and development of schizophrenia (although see (23)), and moderate evidence of increased social anxiety disorder, increased suicidal ideation, and increased episodes of mania in bipolar disorder with regular use (10). In some rare but serious cases, cannabis use could even result in cardiac death and hyperemesis syndrome (7–9). The cost (3), interactions (6), and adverse effects (7–10) of MC should be taken seriously and evidence-based medicine should always be a priority. However, due to the rarity of life-threatening adverse events, it must be noted that even with these potential dangers, MC remains a relatively safe option in the world of pharmaceuticals.

Our time analysis showed that the number of QCs with “partial” fit to NAS-established evidence categories increased significantly from 2017 to 2024. However, the *percentage* of partial QCs decreased. “Partial” was the largest category in both 2017 (44% of QCs) and 2024 (37%). Those within this category partially fit into one or more categories, each varying in level of evidence. For example, 94% of states include some variation of “HIV or AIDS” in their QC list. We classified this QC as partial because the NAS report found that there was limited evidence of effectiveness specifically for “increasing appetite and decreasing weight loss in HIV/AIDS” (10). The NAS report searched for evidence of cannabis’s effect on specific symptoms of diseases, the way scientific research on cannabis is reported. Because lawmakers, not clinicians, are creating states’ QC lists, their lists often simply state diseases, which could encompass a range of pathophysiology, symptoms, and treatments. Another issue that arises is different disease states being lumped together. For example, one Hawaii QC reads “persistent muscle spasms, including those characteristic of multiple sclerosis or Crohn’s Disease.” Although both diseases may generate muscle spasms, those spams are of very different etiologies and involve different organ systems—skeletal muscle versus enteric muscle. The un-scientific nature of creating QCs creates a vague process for certifying providers to work from, rather than a scientific and research-based approach that uses existing nomenclature (e.g. the International Classification of Diseases or the Diagnostic and Statistical Manual of Mental Disorders).

The NAS report is a snapshot of the accumulated evidence of medical science on MC in 2017. As a variety of QCs are investigated, new evidence will emerge for use of MC. The NAS report concluded insufficient evidence for epilepsy, although in 2018 the FDA approved Epidiolex, a cannabidiol used for certain seizure disorders (24). This is the first and only CBD product approved by the FDA. In some cases, more current research supports the findings in the NAS report. For example, chronic pain was rated by the NAS report to be one of few conditions for which MC has substantial evidence of effectiveness (10). In a 2022 systematic review, high THC-to-CBD ratio products were found to have a moderate effect on pain symptoms, and another in the same year found both dronabinol and nabiximols, which are MC products, to have moderate evidence of therapeutic effect for chronic pain (25,26). Conclusions made by the NAS report for other QCs similarly remain true. A 2020 systematic review found “potential” effect of cannabis for PTSD but labeled those studies as small and having “methodological weaknesses” (25,27). Another systematic review found a positive effect of MC for PTSD but could not come to a conclusion on the value of evidence (28). These results uphold the NAS report’s finding that evidence for beneficial effects of MC for PTSD is limited. However, a Canadian study that used a patient self-reported survey to assess MC effects on PTSD found that patients felt an average of 49% better after six weeks of treatment (29). This real-world evidence has the unique benefit that participants in this study were using the range of MC products that are accessible to patients on the market. New information like this should be taken together with the finding of the NAS report and be incorporated into states’ determination of QCs rather than replacing established accumulation of evidence. Overall, while the NAS report (10) reflects evidence at the time it was published, it largely mirrors many of the more recent systematic reviews and meta-analyses, as well as the 2023 Mayo Clinic report on medical marijuana (4,25,30,31).

## Limitations

Based on the nature of the data, it was difficult to create an objective analysis. As discussed, not every QC exactly matches a condition or symptom studied by the NAS report. Therefore, the authors had to make their own decisions about which categories those states fit in to. For example, New Hampshire adds a qualifier to its list that for each QC to count, that condition must also include “at least one of the following: agitation of Alzheimer’s disease, cachexia, chemotherapy-induced anorexia, constant or severe nausea, elevated intraocular pressure, moderate to severe insomnia, moderate to severe vomiting, seizures, severe pain, severe persistent muscle spasms, wasting syndrome.” Of course, each of those extra symptoms could be its own QC with its own level of evidence, making it difficult to separate the listed QC from the extra symptoms required to be included.

Partial QCs were especially difficult, and we would classify the need for a partial category as a limitation of this work. However, the partial category also emphasizes our conclusion that QC list creation, being handled by politicians and not medical practitioners, lacks an objective and scientific process.

## Conclusion

In conclusion, this report provides valuable insight into the disconnect between data and medical practice and may be used to inform future policy decisions. It is possible that states are using other information to guide their QCs, including voter initiatives and public opinion (32), which may explain why the legislation seems to be moving faster than evidence. As new research is completed, those findings should be added to the foundation that the NAS report has provided.

## Conflicts of Interest

The work was completed with software from the National Institute of Environmental Health Sciences [T32-ES007060-31A1]; BJP is supported by the Health Resources Services Administration [D34HP31025] and was (2019-21) part of an osteoarthritis research team supported by Pfizer and Eli Lilly. This work was supported by Ascend Wellness and the Geisinger Academic Clinical Research Center (Grant number 004). They had no role in the design of the study or the publication decision.

A preliminary report of this work was previously published on MedRxiv, 5/10/23. Doi: https://doi.org/10.1101/2023.05.01.23289286

## Ethics

This report was approved by the IRB of Geisinger as exempt.

## Data Availability

All data produced in the present study are available upon reasonable request to the authors.

## Abbreviations

MC: medical cannabis
NAS: National Academy of Sciences
QC: Qualifying Conditions
US: United States

## Notes

### Summary of Updates

Data from 2024 was added and the figures were updated.

## References

1. Rhee TG, Rosenheck RA. Increasing Use of Cannabis for Medical Purposes Among U.S. Residents, 2013-2020. Am J Prev Med. 2023 Sep;65(3):528–33.

2. NORML. Medical Marijuana Laws [Internet]. Available from: https://norml.org/laws/medical-laws/

3. Piper BJ, Beals ML, Abess AT, Nichols SD, Martin MW, Cobb CM, et al. Chronic pain patients’ perspectives of medical cannabis. Pain. 2017 Jul;158(7):1373–9.

4. Solmi M, De Toffol M, Kim JY, Choi MJ, Stubbs B, Thompson T, et al. Balancing risks and benefits of cannabis use: umbrella review of meta-analyses of randomised controlled trials and observational studies. BMJ. 2023 Aug 30;e072348.

5. Department of Health [Internet]. [cited 2024 Mar 3]. Medical Marijuana Assistance Program (MMAP). Available from: https://www.health.pa.gov:443/topics/programs/Medical%20Marijuana/Pages/MMAP.as px

6. Lexicomp: Evidence-Based Drug Referential Content. Lexicomp Drug Interactions: Dronabinol. Available from: www.uptodate.com/drug-interactions/?search=dronabinol&topicId=9394&source=responsive_topic#di-analyze

7. Hartung B, Kauferstein S, Ritz-Timme S, Daldrup T. Sudden unexpected death under acute influence of cannabis. Forensic Sci Int. 2014 Apr;237:e11–3.

8. Patel M, Sathiya Narayanan R, Peela AS. A Case of a Patient With Cannabis Hyperemesis Syndrome Along With Recurrent Nephrolithiasis. Cureus [Internet]. 2023 Apr 5 [cited 2023 Oct 15]; Available from: https://www.cureus.com/articles/142401-a-case-of-a-patient-with-cannabis-hyperemesis-syndrome-along-with-recurrent-nephrolithiasis

9. Schreck B, Wagneur N, Caillet P, Gérardin M, Cholet J, Spadari M, et al. Cannabinoid hyperemesis syndrome: Review of the literature and of cases reported to the French addictovigilance network. Drug Alcohol Depend. 2018 Jan;182:27–32.

10. National Academies of Sciences, Engineering, and Medicine (U.S.), editor. The health effects of cannabis and cannabinoids: the current state of evidence and recommendations for research. Washington, DC: The National Academies Press; 2017. 468 p.

11. Hanson K. How Four States Incorporated Public Health and Cannabis Policy: A Case Study [Internet]. The National Conference of State Legislatures; 2022 Aug. Available from: https://documents.ncsl.org/wwwncsl/Health/NCSL-PH-and-Cannabis-Policy.pdf

12. Ohio Medical Marijuana Control Program [Internet]. [cited 2023 Dec 13]. Available from: https://medicalmarijuana.ohio.gov/faqs

13. 4470 State of Delaware Medical Marijuana Code [Internet]. [cited 2023 Dec 13]. Available from: http://regulations.delaware.gov/AdminCode/title16/Department%20of%20Health%20and%20Social%20Services/Division%20of%20Public%20Health/Health%20Systems%20Protection%20(HSP)/4470.shtml#1058727

14. Hirsch AG, Wright EA, Nordberg CM, DeWalle J, Stains EL, Kennalley AL, et al. Dispensaries and Medical Marijuana Certifications and Indications: Unveiling the Geographic Connections in Pennsylvania, USA. Med Cannabis Cannabinoids. 2024 Feb 20;7(1):34–43.

15. Boehnke KF, Gangopadhyay S, Clauw DJ, Haffajee RL. Qualifying Conditions of Medical Cannabis License Holders in the United States. Health Aff Proj Hope. 2019 Feb;38(2):295–302.

16. Wayback Machine. Internet archive: digital library of free & borrowable books, movies, music & wayback machine [Internet]. Available from: https://archive.org/

17. GraphPad Prism. (9.3.1.).

18. About Us - American Glaucoma Society [Internet]. [cited 2024 Feb 9]. Available from: https://www.americanglaucomasociety.net/about/statements

19. Parkinson’s Foundation Consensus Statement on the Use of Medical Cannabis for Parkinson’s Disease [Internet]. Parkinson’s Foundation; 2020. Available from: https://www.parkinson.org/sites/default/files/documents/medical_cannabis_statement_finalv2_5.pdf

20. Boehnke KF, Sinclair R, Gordon F, Hosanagar A, Roehler DR, Smith T, et al. Trends in U.S. Medical Cannabis Registrations, Authorizing Clinicians, and Reasons for Use From 2020 to 2022. Ann Intern Med. 2024 Apr 9;

21. Driving and traveling | Colorado Cannabis [Internet]. [cited 2024 Apr 15]. Available from: https://cannabis.colorado.gov/legal-marijuana-use/driving-and-traveling

22. Field Sobriety Tests and THC Levels Unreliable Indicators of Marijuana Intoxication | National Institute of Justice [Internet]. [cited 2024 Apr 15]. Available from: https://nij.ojp.gov/topics/articles/field-sobriety-tests-and-thc-levels-unreliable-indicators-marijuana-intoxication

23. Elser H, Humphreys K, Kiang MV, Mehta S, Yoon JH, Faustman WO, et al. State Cannabis Legalization and Psychosis-Related Health Care Utilization. JAMA Netw Open. 2023 Jan 3;6(1):e2252689.

24. Commissioner O of the. FDA. FDA; 2018 [cited 2024 Apr 15]. FDA Approves First Drug Comprised of an Active Ingredient Derived from Marijuana to Treat Rare, Severe Forms of Epilepsy. Available from: https://www.fda.gov/news-events/press-announcements/fda-approves-first-drug-comprised-active-ingredient-derived-marijuana-treat-rare-severe-forms

25. Bilbao A, Spanagel R. Medical cannabinoids: a pharmacology-based systematic review and meta-analysis for all relevant medical indications. BMC Med. 2022 Aug 19;20(1):259.

26. McDonagh MS, Morasco BJ, Wagner J, Ahmed AY, Fu R, Kansagara D, et al. Cannabis-Based Products for Chronic Pain: A Systematic Review. Ann Intern Med. 2022 Aug;175(8):1143–53.

27. McKee KA, Hmidan A, Crocker CE, Lam RW, Meyer JH, Crockford D, et al. Potential therapeutic benefits of cannabinoid products in adult psychiatric disorders: A systematic review and meta-analysis of randomised controlled trials. J Psychiatr Res. 2021 Aug;140:267–81.

28. Sarris J, Sinclair J, Karamacoska D, Davidson M, Firth J. Medicinal cannabis for psychiatric disorders: a clinically-focused systematic review. BMC Psychiatry. 2020 Dec;20(1):24.

29. Cahill SP, Lunn SE, Diaz P, Page JE. Evaluation of Patient Reported Safety and Efficacy of Cannabis From a Survey of Medical Cannabis Patients in Canada. Front Public Health. 2021 May 20;9:626853.

30. The Editors of Mayo Clinic. Medical Marijuana: the science and the benefits. Meredith Operations Corporation; 2023.

31. Allan GM, Finley CR, Ton J, Perry D, Ramji J, Crawford K, et al. Systematic review of systematic reviews for medical cannabinoids: Pain, nausea and vomiting, spasticity, and harms. Can Fam Physician Med Fam Can. 2018 Feb;64(2):e78–94.

32. Schlag AK, Zafar RR, Lynskey MT, Athanasiou-Fragkouli A, Phillips LD, Nutt DJ. The value of real world evidence: The case of medical cannabis. Front Psychiatry. 2022 Nov 3;13:1027159.

